# A Comparison between Evidence-Generated Transtibial Sockets and Conventional Computer-Aided Designs, from the Patient’s Perspective

**DOI:** 10.1101/2024.09.17.24312762

**Authors:** Florence Mbithi, Maggie Donovan-Hall, Jenny Bramley, Joshua Steer, Charalambos Rossides, Peter Worsley, Chantel Ostler, Cheryl Metcalf, Dominic Hannett, Caroline Ward, Jack Kitchen, Sioned Steventon, Katy McIntosh, Shigong Guo, Helen Harvey, David Henderson Slater, Vijay Kolli, Alex Dickinson

## Abstract

**Objective:** Personalised prosthetic socket design depends upon skilled prosthetists who aim to balance functional human-prosthesis coupling with safe, comfortable load transmission to skin and soft tissues. This study’s objective was to assess the comfort of sockets generated from past computer aided socket design records.

**Design:** A crossover non-inferiority trial with embedded qualitative interview study.

**Setting:** Three United Kingdom National Health Service clinics.

**Participants:** Seventeen people with nineteen transtibial amputations.

**Intervention:** Evidence-Generated sockets and conventional clinician-led computer aided (Control) designs

**Main Measures:** Socket Comfort Score and semi-structured interview.

**Results:** Evidence-Generated sockets had no statistically-significant difference in comfort compared to clinician-led Control sockets (p=0.38, effect size=0.08), but a lower socket comfort score variability across the group. Analysis of interviews revealed themes around fitting session experiences, similarities and differences between the Evidence-Generated and Control sockets, and residual limb factors impacting perceptions of socket comfort. These provided insights into the participants’ experience of the study and the value of expert prosthetist input in socket design.

**Conclusions:** Evidence-Generated sockets demonstrated noninferiority to conventional clinical computer aided design practice in terms of socket comfort. Both quantitative and qualitative results indicated how clinician input remains essential and is valued by prosthesis users. Work is underway to incorporate the evidence-generated sockets into computer aided design software such that they can act as a digital starting point for modification by expert clinicians at fitting, potentially reducing time spent on basic design, enabling prosthetists to focus on more highly-skilled customisation and co-design with their patients.

## Introduction

The lower limb prosthetics community has worked since the 1980s towards computer aided design and manufacturing (CAD/CAM) technologies to support prosthetic socket design and fabrication workflows. These technologies were proposed to offer evidence-based decision support (1). Computer Aided Socket Design (CASD) has been described as offering significant potential benefits over conventional plaster of Paris approaches [2]. CASD can reduce the manual work time and occupational exposure to plaster. This could allow clinicians to spend a greater proportion of their time in direct patient interactions, which might enhance patient engagement and facilitate shared decision-making [2]. Early CAD/CAM results were comparable to traditionally-produced sockets but took longer and needed more adjustments (1–4). However, over the learning curve, today clinicians using CAD/CAM can achieve clinical results that are comparable whilst saving time and delivering a better quality of life (QoL) outcome (5–7).

The digital approach also generates a perpetual, quantitative design record, whereas the plaster design is destroyed upon socket fabrication. A digital record has value for education, peer support on complex cases and, if implemented for clinical decision support, further time savings to focus on the most highly skilled and value-added parts of socket personalisation (3,8). However, despite proposals since the 1980s to augment CAD/CAM in these ways (9) it appears that the full potential of the benefits offered through these rich data sets to improve the prosthetic rehabilitation process have not been fully exploited.

In particular, data might be used for generating evidence-based socket design recommendations. Since Dean and Saunders in 1985 (10), clinical innovators have considered an alternative to performing transtibial CASD in a manner analogous to manual work with plaster, whereby a user could apply averaged rectifications from prior designs as an ‘overlay’, or select from a database of different size and shape ‘primitive’ or ‘template’ sockets, and scaling and editing them to fit to a newly presenting patient’s residual limb shape. This has been demonstrated for transtibial (3,8,11,12) and transfemoral socket designs (13,14), and in orthotics for scoliosis braces (15,16). Recent publications present alternative methods of data-informed socket design including Fuzzy Logic or Inference to map linguistic descriptions of lower limb socket design approaches and patient descriptors, for application to new people (17,18), and optimisation for automating transradial socket design (19). However, these studies are only demonstrated in a research setting and not yet clinically applied.

Anecdotally, most CASD software packages in clinical use define templates as structured design workflows or sequences to guide users to apply their own choice of rectifications and gross design features.

The application of such methods would benefit from systematic study of rectification design practice, though this is limited in the scientific literature (20) since the AFMA project (11). Recently researchers have begun to leverage high resolution 3D scan and CASD data to investigate rectification sizes in TT and TR design (21–23), and most recently probabilistic methods have been used to derive insights into TT socket design strategy through the associations between design features (24). Building upon those insights, the present study aims to evaluate an evidence-generated socket concept developed by Radii Devices Ltd. and University of Southampton, with UK service provider Opcare Ltd providing data and expert clinical design insight. The objective was to compare the evidence-generated socket design approach to clinician-led CAD/CAM, using socket comfort outcome measures and capturing the patients’ experiences through qualitative interviews.

## Methods

This study is reported using the STROBE cross sectional reporting guidelines (25).

### Patient and Participant Involvement and Engagement (PPIE)

The study research question was informed by PPIE group discussions (26), which highlighted how socket comfort is paramount but difficult to achieve, and that delays between assessment and device provision can impair fitting. PPIE contributors also expressed support for sharing knowledge between prosthetics centres to enable service improvement. Collaboration with the Alex Lewis Trust during the study development reinforced this, and provided review and feedback of the study design, recruitment posters, participant information sheets and consent forms. PPIE contributors are also actively involved in disseminating the study findings to patients.

### Study Design

An Evidence-Generated (EG) transtibial check socket was compared to a control socket designed by a prosthetist using Tracer CAD/CAM software (Ohio Willowwood Co, USA) from the same residual limb 3D scan (Figure 1). The study used a single-blind, crossover design to assess comfort at socket fitting, with an embedded qualitative study of semi-structured interviews. Quantitative and qualitative data were collected and analysed independently, and interpreted together. This study design was selected to capture the patient’s comparative experience of socket fitting from quantitative and qualitative perspectives, developed from a foundational CAD/CAM vs. traditional socket comparison study (5) and with recent precedent (27). This study design was chosen because:

- new digital design and fabrication technologies are often considered in small scale or low technology readiness level (TRL) trials in the scientific literature but are often not tested in power-sized, blinded or controlled trials, with consideration of qualitative service user experience alongside quantitative outcome measures (20); and
- it is important to understand the patient’s perceived experience. This may relate to socket fit and comfort for which detailed, open descriptions offer more nuanced insights than a simple socket comfort score (28). It may also relate to the user’s perspective of device design and fabrication, to promote shared-decision-making (29) and its potential benefits regarding patient engagement in and understanding of care.

**Figure 1.**
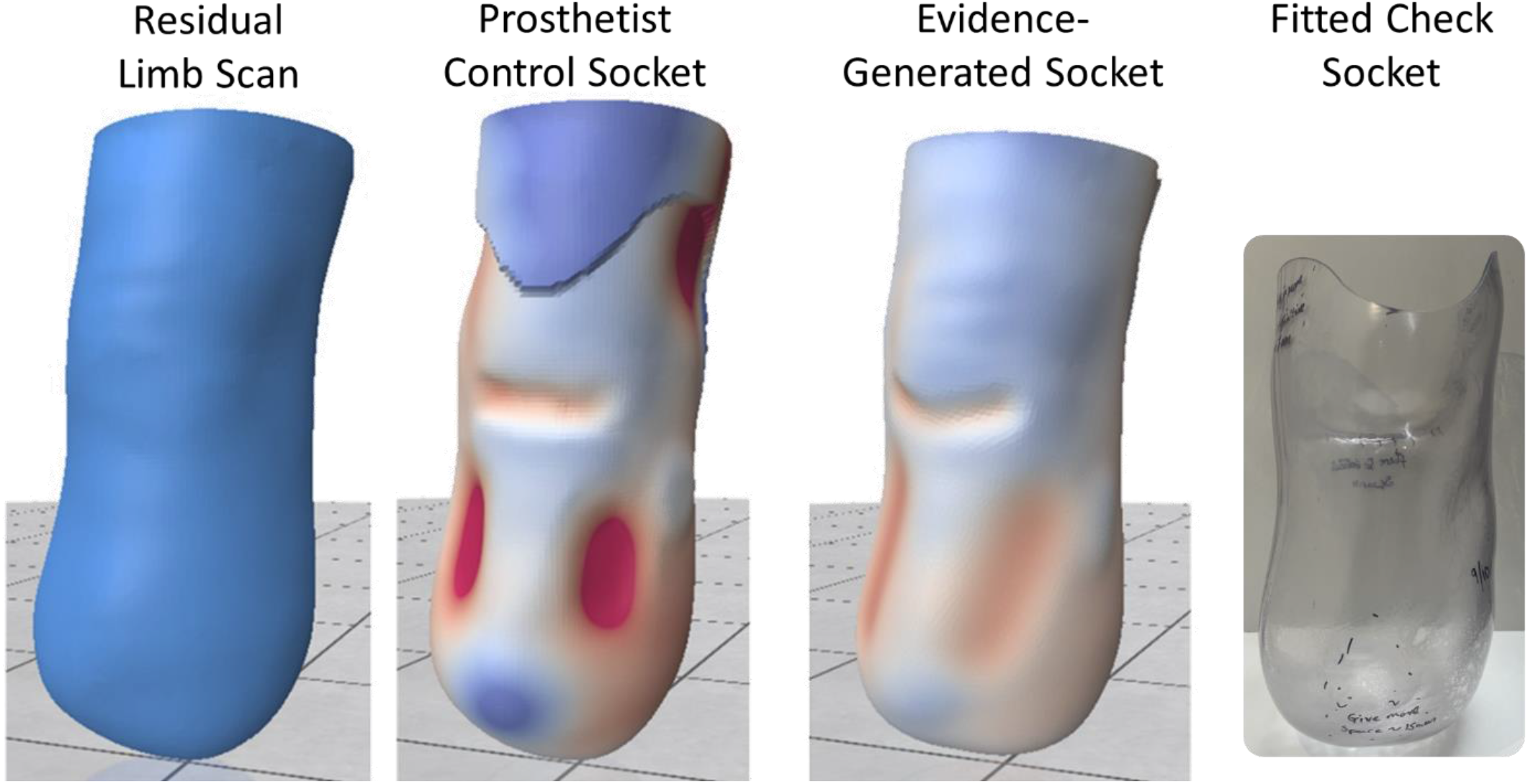
from left to right, for an exemplar participant: the residual limb scan over a sock; the prosthetist’s socket rectification design; the evidence-generated design prior to adding brimline; and a fitted check socket. In rectification design maps, red colour represents a carve or press-fit between socket and limb, white represents exact fit, and blue represents buildup or limb-socket gap.

As such the work aligns with recently published perspectives on ethical considerations for development and clinical translation of prosthetic technologies (30).

### Study Approvals

Ethical approval was granted by Institutional (ERGO 76033.A3) and national review boards (IRAS 313408 / HRA REC 22/YH/0215). Inclusion criteria included people aged 18 and over, with transtibial amputation, deemed ready for a new prosthetic socket by their prosthetist in their usual clinical pathway, and willing and able to tolerate trialling two sockets at a fitting session. Convenience sampling was used to identify participants at three UK prosthetic rehabilitation centres.

### Evidence-Generated Socket Design Method

A socket generation method based upon evidence of expert practice was applied, building upon previous work (24). To review briefly, a dataset of 163 transtibial residual limb 3D scans and corresponding socket designs was accessed in .aop format including labelled landmarks. The corresponding patient characteristics such as gender, age, time since amputation, and reason for amputation were also collated. A Statistical Shape Model (SSM) was generated to describe the residual limb shape and size with a minimal number of dimensions, corresponding to principal ‘modes’ of variation. Socket design features of local rectifications (patella tendon bar carve, paratibial carves, fibula head build, distal end build, distal tibia build, anterior tibia build, supracondylar carves) and gross volume change were extracted from the dataset. Bayesian inference was applied to analyse the probabilistic association between patient characteristics and principal modes of limb shape variation (inputs), and the extracted design features for the 163 limb-socket pairs. Finally, the same inputs were obtained for the study participants, and check sockets were created by automatically applying modifications to their landmarked residual limb 3D scans, which were predicted using the statistical model.

### Quantitative study

The evidence-generated socket design was generated automatically except for the brim line, which the fabrication technician was requested to apply in the same location as the control, prosthetist-designed socket. The prosthetist then worked through their standard fitting and assessment procedure for both sockets, including a Socket Comfort Score. At the fitting appointment, the prosthetist chose which socket was trialled first, and the patient participant was blinded to which socket was prosthetist-designed or evidence-generated. At the end of the session, the patient participant and their clinician were allowed free choice over which check socket design was to be used for the definitive prosthesis, on the basis of their mutual agreement without any intervention from the researchers.

A power calculation indicated a sample size of 19 was required to test non-inferiority in a crossover trial (power: 0.9, significance level: 0.05, Mean Difference: 0, non-inferiority limit: 1.21 (31), and population SCS standard deviation 1.2 (21)). A Shapiro-Wilk test indicated the control socket SCS data were not normally distributed (p=0.01), so non-parametric descriptive statistics were calculated with confidence intervals using the Bootstrap method (32), and the paired Wilcoxon Signed-Rank test was used to assess the statistical significance of difference between the sockets.

### Embedded Qualitative study

To capture participants’ views and experiences of in-clinic fitting of two sockets designed in different ways, and general usability of their new prosthetic socket, two semi-structured interviews were carried out. The first at the time of the socket fitting and second one-month afterwards (Appendix). Interviews were audio-recorded, anonymised, and transcribed verbatim by a professional transcribing company. A member of the research team (FMM) checked a sample of transcripts against the audio recordings for accuracy. The transcripts were analysed using thematic analysis, which provided a flexible approach to ascertain a clear understanding of the comparability of socket comfort and views of the processes (33). The thematic analysis approach consisted of 1) familiarisation with the data, 2) coding by hand and using NVIVO software (QSR International, Melbourne, Australia), 3) generating initial themes, and 4) reviewing and finalising final themes to capture consistent patterns within the data and key meaning relevant to the research questions (33). One team member (MDH) led all stages of the analysis. Two other members coded different sections of the data and together agreed on the final themes (FMM, JLB), providing verification, and allowing for a range of interpretations.

## Results

Seventeen recruited participants with nineteen residual limbs completed the study between March and November 2023 (Table 1). Six prosthetists designed control sockets for between one and nine participants. Approval was initially granted to compare transparent check sockets, but due to low recruitment a study protocol amendment was approved to compare the evidence-generated check socket to a definitive prosthetist-designed socket. Therefore, single-blinding and offering choice of socket for definitive fabrication could not apply to three participants for whom the evidence generated check socket was compared to a definitive socket control.

**Table 1:** Description of Participants.

|  |  |
| --- | --- |
| Side (n=19) | 8 Left / 11 Right |
| Sex (n=17) | 1 Female / 16 Male |
| Age (yrs, n=17) | Median 66, min 30, max 84 |
| Time since amputation (yrs, n=19) | Median 4.0, min 0.7, max 16.1 |
| Reason for amputation (n=19) | 6 Infection / diabetic foot<br>4 Dysvascularity<br>3 Trauma<br>3 Ischaemia / CLTI<br>1 Sepsis<br>1 Peripheral Neuropathy<br>1 Chronic osteomyelitis secondary to trauma |
| Reason(s) for new socket (n=19) | 12 socket too large / limb shrinkage<br>3 requested new suspension method<br>2 socket too small / tight<br>1 alignment incorrect<br>3 N/R |
| Participant Activity Level (n=17)<br>A1: In house walking or transfers only<br>A2: Walking on flat ground only<br>A3: Normal everyday walking<br>A4: Additional high impact/energy activities | 1 A1<br>6 A2<br>8 A3<br>0 A4<br>2 N/R |

Notes: N/R denotes 'not reported'. CLTI: critical limb-threatening ischemia.

### Quantitative findings

The unmodified evidence-generated sockets had no statistically significant difference in socket comfort scores compared to prosthetist-designed control sockets (median SCS 8.6 for EGs and 8.8 for controls; p=0.376 and effect size 0.08 in the paired significance test, Figure 2). Lower variability in SCS was observed across the study group for the EG sockets than the control sockets (95% confidence intervals 8.0-9.0 and 7.5-9.5, respectively). Nine of the nineteen EG sockets were given a comfort score within the noninferiority limit (1.21) of the control. Six EG sockets were rated as more comfortable than the control, by between 2 and 3.5 points. Four EG sockets demonstrated lower comfort than the control, by 1.5 to 2.5 points.

**Figure 2.**
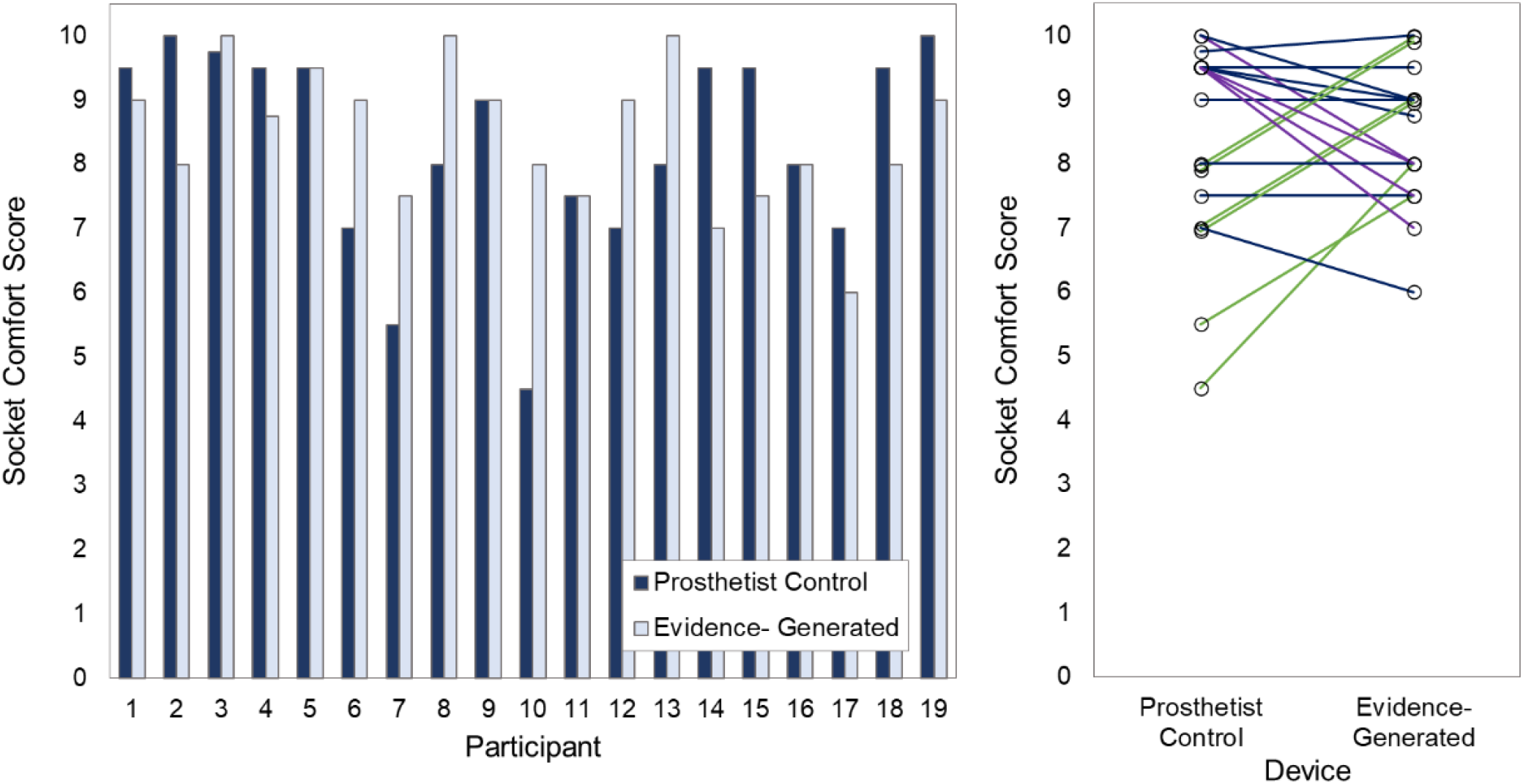
noninferiority crossover study results comparing Socket Comfort Score between the prosthetist control and evidence-generated sockets, for 19 fittings across 17 participants (left) and paired comparison chart (right). Colour coding denotes higher (green), the same (blue) or lower (purple) SCS for EG than prosthetist control devices, within the noninferiority threshold of 1.21 points.

### Qualitative findings

Due to the significant impact and diverse experiences related to socket comfort, thematic analysis identified several broader themes that were beyond this paper’s objective. Focus is therefore limited to specific themes that offer insights into the participants’ views and experiences relating to the sockets produced by the two design approaches (Table 2).

**Table 2:** Themes identified in semi-structured interviews relating to comparison between comfort of the fitted sockets and reflections on the socket fitting process:

|  |  |
| --- | --- |
| Theme 1 | Experiences of the fitting session |
| Theme 2 | Similarity between the EG and control sockets |
| Theme 3 | Differences between the EG and control sockets |
| Theme 4 | Residual limb factors impacting on perceptions of socket comfort |

#### Theme 1: Experiences of the fitting session

When discussing the fitting session, participants reflected on several aspects that they felt impacted on their experiences. Several participants described feeling that it went “*well”, “fine”* or *“ok”*, which illustrated that a process involving two different sockets was not a particular issue. For example, participant 10 (male, in their 50s, amputation following trauma) discussed their experience in terms of the output of the session: “*I got something out of it, it looks like this time if you know what I mean. At the end of this fitting there was something that looked like it was ready for me”*. Other participants discussed how they felt about comparing the two sockets and commented on how they did not know who designed either of the two sockets, for example “*I didn’t know who designed and who produced each socket”* (participant 1, male, in their 80s, infection).

Other participants discussed their experience in terms of their expectations of addressing their specific socket fit issues relating to their residual limb. For example, “*The only thing I’ve got hope for there is that the bottom of my stump shrinks a lot more before it gets thinner and thinner because that’s the only thing that’s stopping me having a smaller prosthetic is that stump. It’s where they added all the bits on”* (participant 10, male, in their 50s, amputation following trauma).

#### Theme 2: Similarity between the evidence-generated and control sockets

Several participants described not feeling any difference in the level of comfort between the sockets regardless of which design process was used. For some participants, this appeared to be related to feeling that both sockets were equally comfortable, with descriptions of how they found it difficult to decide if one was more comfortable than the other: *“As far as comfort there was very little difference in it really to be honest as far as I could judge at the time”* (participant 19, male, in their 70s, amputation due to ischemia). Some participants compared the comfort of the two new sockets to their previous socket. For example: “*I feel a lot more comfortable really than the old one”* (participant 1, male, in their 80s, infection).

This similarity in the level of comfort between the two new sockets was described by some participants in relation to both sockets feeling equally *“firm”, “stable”, “both fitting well”* and both sockets feeling very *“natural”*, as described by participant 4 (male, in their 30s, elective amputation following trauma): “*Absolutely fine. They were both very, very close to being completely natural”*. However, for other participants, the similarity between the sockets appeared to be associated with feeling similar levels of pain: “*walking with them as I was walking there was the same amount of pain”* (participant 11, female, in their 80s, peripheral vascular disease).

#### Theme 3: Differences between the evidence-generated and control sockets

Other participants noticed significant differences in the comfort of the two sockets, from positive and negative perspectives. They mentioned issues such as pressure points, discomfort at the back of the leg, and feelings of tightness. For example: “*One felt really tight. I like it tight when I get a new socket because it lasts longer. One felt a lot more comfortable at the front where my bone is closer to the skin”* (participant 12, male, in their 40s, amputation due to chronic ulcers).

Differences in the size of the sockets also affected comfort. Participant 4 (male, in their 30s, elective amputation post trauma) explained, “*The taller one was a bit more uncomfortable due to how high it was. When you bend your leg down, it hits the back of your leg*.” Some participants felt that the height of the socket made it feel more natural. These differences impacted participants’ walking, with one socket feeling easier to walk in than the other: “*One socket was absolutely perfect. The other one had different movement, slightly different, but it seemed like you walked quicker with it. It was a nice, easy movement”* (participant 8, male, in their 70s, ischaemic amputation).

#### Theme 4: Residual limb factors impacting on perceptions of socket comfort

While comparing the two sockets, participants discussed sensitive areas that affected their comfort, such as spots on the crest of the shin and other specific sore areas. These issues were often ongoing and considered by the prosthetist during socket design: *“I’ve always had this issue. We’ve had to shave out a bit of the socket to ease the pressure. But I think it’s just the shape of my stump, and it’s something that will always be an issue and I’m always aware of it”* (participant 13, male, in their 30s, elective amputation post trauma). Participant 16/17 (male, in their 70s, diabetic amputations) reflected on the adjustment after the fitting process, saying “*I don’t know if the second one on the right whether it needed any more adjustment. Maybe a little bit but I would say they are fairly comfortable*.*”*

Other factors also not necessarily directly related to the design process appeared to impact on the participants’ views of which of the two new sockets were more comfortable. For example, participant 6 (male, in their 50s, dysvascular amputation) described how their residual limb had shrunk since their last prosthetic fitting and this led to the new socket feeling more comfortable: *“But it does feel better and that clear one that I had in my old socket … had more movement because the stump had shrunk so much. So, it’s just a case of getting used to having this because it’s slightly smaller and it’s hugging the stump. Perhaps where I had the freedom before where it moves about a lot more … I’ve got to get used to the new feelings of it*.*”*

## Discussion

This study demonstrated noninferiority of the evidence-generated socket design method in comparison to conventional clinician-led computer aided socket design at initial fitting. Socket comfort score indicated similar comfort on average, and reduced variability across the cohort, for the evidence-generated sockets. Participant interviews confirmed this assessment and added a more detailed understanding of socket satisfaction.

Thematic analysis also revealed the patients’ detailed understanding of the nuances of their prosthesis design and its fit, and demonstrated the importance of ensuring a trained, experienced clinician directs the application of technology-enhanced socket design processes. The comfort results indicated non-inferiority of the evidence-generated sockets’ fundamental design without any personalised clinician input. However, the embedded qualitative study provided evidence to support clinical usage of evidence-generated sockets with an expert human in the loop to make design decisions in response to patients’ individual and complex needs (11,21,34). That might include local design modifications in response to vulnerable sites on that individual’s residual limb or accommodating the patient’s preferences for tightness of fit. This was identified for the single participant whose evidence-generated sockets was rated with socket comfort score below seven (Participant 16/17, male, in their 70s, diabetic amputation), for whom the score of seven was attributed to an unusual tissue sensitivity at a supracondylar site, where the control socket featured a local modification. The same point may explain the observed trend where all four evidence-generated sockets that were rated as less comfortable than the control were cases where the control scored a socket comfort score of nine to ten, where such personalisation has evidently been successful. Patient-specific local areas of sensitivity and corresponding design changes inherently cannot be generalised, so this justifies the importance of an expert in the design process, who retains clinical responsibility.

The complementary evidence provided by this parallel quantitative-qualitative approach demonstrates the value of exploring the participants’ experience more deeply through interviewing, to enhance what can be learned from objective measurement (35). However, this approach is somewhat unusual in the assessment of prosthetics technologies (27). The study involved a large interdisciplinary team which enabled separation of trial socket design, clinical assessment, interviewing, data analysis and interpretation, in an attempt to minimise potential researcher biases. The combined synthesis of the socket comfort score and interviews revealed the participants’ detailed understanding of the influence of residual limb shape and corresponding socket design upon their comfort and function, illustrative of the value of excellent prosthetist-patient communication. These technologies may offer an opportunity to improve patient experience through understanding of their care (36) or even shared decision making (37). The use of an evidence-generated socket design process may enable greater focus on the higher value-added aspects of personalisation, and might be easier to perform in front of the patient (21). Most notably this observation justifies ongoing work with clinician stakeholders, for example to indicate specific areas in which human input is particularly important for fitting evidence-generated sockets to newly presenting individuals, and in development of software interfaces, building upon methods reported by Ngan et al (38).

The study included a relatively diverse group of patients accessing prosthetic rehabilitation services except for gender, where the recruited participants were predominantly male (16/17) (39), which may impact the study’s generalisability. Women are less likely to have a major amputation and to be successfully fitted with a prosthesis (40), enter prosthetic rehabilitation later (41), and are under-represented in research cohorts (42). It is not clear why the present study’s convenience sampling resulted in an imbalance, but this illustrates the need to ensure that further studies include more diverse gender representation, as the experiences of men and women may be different. However, the distribution of randomly sampled people whose socket designs were used to train the socket design evidence model (24) was representative of the population of people with transtibial amputations, and external validity is supported by their diverse range of ages, reasons for limb absence, activity levels and time since amputation. The study also has limitations in its single timepoint and outcome measure, where previous studies comparing computer aided design and manufacturing to plaster design and fabrication considered quality of life (7) and more extensive assessments of comfort, fit, cosmesis, weight and function alongside clinical workload and productivity measures (8). The study cannot indicate how socket comfort would develop long-term, and it may be preferable to use the Expanded Socket Comfort Score for Best, Worst and Average Comfort over a longer period (43). However, this study replicates the clinical assessment of trial and definitive sockets in UK National Health Service settings, and provides additional insight since participants could compare two socket options side-by-side and select their preference, with interviews proceeding a month after fitting. Finally, the study was not formally randomised and prosthetists generally fitted their own-design socket first, meaning a carryover effect cannot be excluded.

Overall, the study findings support wider clinical use of evidence-generated transtibial sockets, but also demonstrates the importance of delivering this technology in a way that facilitates prosthetist input in tailoring the design to their individual patients’ needs and provides maximal opportunity for their communication. As such, we may take the advantages offered by technology whilst retaining or ideally enhancing trust through human-centred prosthetic rehabilitation.

### Clinical Messages

- Evidence-generated transtibial prosthetic socket designs compared well to devices produced by conventional CAD/CAM clinical practice, in terms of comfort at socket fitting and patient feedback in semi-structured interviews.
- Qualitative feedback confirmed that clinician input remains essential, to incorporate patient-specific socket design details in response to local sites of sensitivity or tissue vulnerability, or preference: details that cannot be generalised.
- Work is underway to incorporate evidence-generated socket designs into CAD/CAM software such that they can be modified at fitting, enhancing evidence-based practice as a support tool to help qualified clinicians to leverage their experience and skill and enable co-design with the prosthesis user.

## Acknowledgments

We acknowledge our study participants, patient advisor the Alex Lewis Trust, and kind support from the

University of Southampton’s Institute for Life Sciences.

## Ethical Approval and Informed Consent Statements

The study was conducted according to the guidelines of the Declaration of Helsinki, and was granted approval by Institutional (ERGO 76033.A3) and UK national review boards (IRAS 313408 / HRA REC 22/YH/0215). All participants provided written, informed consent and the presented data are selected to avoid identification. The study is listed on the UK Health Research Authority (HRA) website at https://www.hra.nhs.uk/planning-and-improving-research/application-summaries/research-summaries/feasibility-study-on-using-smart-templates-for-socket-fitting-v2/ and the protocol is registered at the clinicaltrials.gov Protocol Registration and Results System at https://clinicaltrials.gov/study/NCT06597266.

## Data Availability Statement

The dataset supporting the conclusions of this article is available in the University of Southampton repository, https://doi.org/10.5258/SOTON/D3205.

## Appendix: Semi-Structured Interview Guide

### Introduction

- *Briefly explain aims of study*
- *Briefly explain purpose of interview to understand their views and experience of being fitted with the two sockets designed in different ways*
- *We are interested in their own views and experiences*
- *There are no right and wrong answers*
- *Explain what will be done with the data collected*
- *Explain reason for blinding and ask whether they know which socket was which*
- *Do they have any questions?*

### Semi-structured interview questions

- How did you find/can you tell me about your experience of today’s socket fitting session?
  ∘ What do you think went?
  ∘ What didn’t go well?
- How did the two sockets you tried on feel?
  ∘ Were they similar/different?
  ∘ Were certain aspects comfortable/uncomfortable?
  ∘ Could you show me where, on a limb image?
  ∘ How stable did the sockets feel? Any differences?
  ∘ How confident would you feel using the socket? Was there any difference between the two regarding how confident you felt?
  ∘ How well do you feel you would be able to do the activities you want? Any differences between the sockets?
- Can you tell me about you overall experience of getting/using a socket so far?
  ∘ Expectations
  ∘ Goals
  ∘ Positives/frustrations
  ∘ Liners and socks used
  ∘ Do you have any areas of no sensation on your residual limb?
  ∘ Do you have any areas that often feel uncomfortable/comfortable?
  ∘ Do you do anything to make your socket more comfortable or functional to use?
- Is there anything else you would like to share on socket comfort and fit?

